# Spatiotemporal and meteorological trends in dengue transmission in the Dominican Republic, 2015-2019

**DOI:** 10.1101/2023.01.05.23284205

**Authors:** Michael A. Robert, Helena Sofia Rodrigues, Demian Herrera, Juan de Mata Donado Campos, Fernando Morilla, Javier Del Águila Mejía, María Elena Guardado, Ronald Skewes, Manuel Colomé-Hidalgo

## Abstract

Dengue has broadened its global distribution substantially in the past two decades, and many endemic areas are experiencing increases in incidence. The Dominican Republic recently experienced its two largest outbreaks to date with 16,836 reported cases in 2015 and 20,123 reported cases in 2019. With this upward trend in dengue transmission, developing tools to better prepare healthcare systems and mosquito control agencies is of critical importance. Before such tools can be developed, however, we must first better understand potential drivers of dengue transmission. To that end, we focus in this paper on determining trends between climate variables and dengue transmission with an emphasis on eight provinces and the capital city of the Dominican Republic in the period 2015-2019. We present summary statistics for dengue cases, temperature, precipitation, and relative humidity in this period, and we conduct an analysis of correlated lags between climate variables and dengue cases as well as correlated lags among dengue cases in each of the nine locations. We find that the southwestern province of Barahona had the largest dengue incidence in both 2015 and 2019. Among all climate variables considered, lags between temperature variables and dengue cases were the most highly correlated. We found that most locations had significant correlations at lags of zero weeks; however, both Barahona and the northern province of Monte Cristi had significantly correlated lags with other provinces at up to eight weeks. These results can be used to improve predictive models of dengue transmission in the country.

## 1. Introduction

Global incidence of dengue fever has increased substantially in recent decades, with the range of dengue expanding from only nine countries before 1970 to at least 129 countries today (1–4). In addition to its rapid global spread, dengue outbreaks in endemic regions are resulting in increasingly larger numbers of cases and contributing to a growing burden on public health systems. Today, it is estimated that over 390 million people are at risk of contracting dengue [5]. Dengue is primarily distributed across regions of the world with tropical and subtropical climates, although in the past two decades, dengue cases have occurred with greater frequency in temperate zones as well [6–9]. In 2021, 1,254,648 cases and 436 deaths were reported in the Americas [10]. According to the Pan American Health Organization (PAHO) the countries in the Caribbean reporting the most cases of dengue between 2014-2021 are Dominican Republic, Martinique, Guadeloupe, French Guiana, and Cuba, with Dominican Republic having 60% more cases in that time as the country reporting the second highest number [10]. Dominican Republic also reports the highest number of severe dengue cases and deaths in the Caribbean [10]. In 2019, Dominican Republic experienced its largest outbreak to date with a 1,145% increase in cases from 2018 [11]. The cumulative incidence in 2019 was 194.85 cases per 100,000 people, which is a 142% increase from the average incidence between 2005-2014 [12, 13].

With outbreaks becoming increasingly severe in Dominican Republic and other regions, it is more imperative than ever to understand drivers of epidemic dengue. Potential drivers of global spread of dengue include increases in urbanization, more frequent global travel, and changes in temperature and precipitation [14–16]. At local scales, transmission of dengue can also be a function of socioeconomic and demographic characteristics, connectivity to other regions, human behavior, volume of tourism, and rates of migration [14–18]. Many of these variables play an important role in developing and sustaining an environment that is suitable for the vectors of dengue, *Aedes aegypti* and *Aedes albopictus*, which in turn amplifies risk of transmission [19–21].

Because there is an inherent delay in between human cases of dengue resulting from the intermediate mosquito host and a serial interval of 15-17 days, early detection of new dengue outbreaks can be complicated [22, 23]. In the last decade, efforts have been made to improve early detection of dengue outbreaks by improving surveillance and warning [22, 24–26]. These early warning systems are mathematical and statistical models that integrate data to provide predictions for changes in dengue transmission that may indicate outbreaks. Chief among these data are climate variables such as temperature, precipitation, or humidity which are all positively correlated with *Aedes* mosquito populations and dengue transmission [22]. However, before such early warning systems can be developed, relationships between climate variables and dengue cases must be explored to determine which climate variables are most important to local and regional dengue transmission.

In this work, we analyze dengue activity in Dominican Republic between 2015 and 2019 and explore relationships between climate variables and dengue cases. We present descriptive analysis of each data set used in the study along with analysis of correlations in lags between variables. We further investigate correlations in lags between provinces in Dominican Republic to understand potential movement of dengue throughout the country. The work presented herein provides a foundation on which statistical and mathematical models can be constructed to further study drivers of previous outbreaks and to predict future outbreaks.

## 2. Material and Methods

### 2.1 Study site

This study was conducted in Dominican Republic, a Caribbean country that occupies the eastern two-thirds of the Island of Hispaniola. The 2019 estimate of the population size of the Dominican Republic is 10,448,499 people [27]. The country is divided geopolitically into 31 provinces and National District (the capital city) [28].

Dominican Republic has perhaps the most diverse climate of all of the Caribbean islands because of the presence of high mountains and abundant coastal regions [29]. Much of the country, however, has a tropical climate with mean annual temperatures ranging between 22-31 °C [29–32]. Rainy seasons vary geographically with the northern part of the country experiencing heavier rain from November to January and much of the rest of the country having its rainy season May-November. Mean annual precipitation in the country ranges between 400mm in the southwest to more than 2200 mm in the mountain regions [29]. The majority of southern coastal regions experience a mean rainfall of around 1000mm while northern coasts typically have a higher mean annual rainfall of 1600mm or more [29].

In this work, we focus on nine provinces throughout the country. These nine were chosen because they are the provinces for which we were able to obtain both meteorological and epidemiological data between 2015 and 2019. The nine provinces included in this study are Barahona, La Altagracia, La Romana, Monte Cristi, Puerto Plata, Samaná, Santiago, Santo Domingo, and Distrito Nacional (Figure 1). The provinces cover each of the three major regions of the country: North (Monte Cristi, Puerto Plata, Santiago, Samaná), South (Barahona), and East/Southeast (Santo Domingo, Distrito Nacional, La Romana, La Altagracia). The nine provinces include 6,295,775 people (2019 estimate), representing 60.78% of the total population of the country.

**Figure 1.**
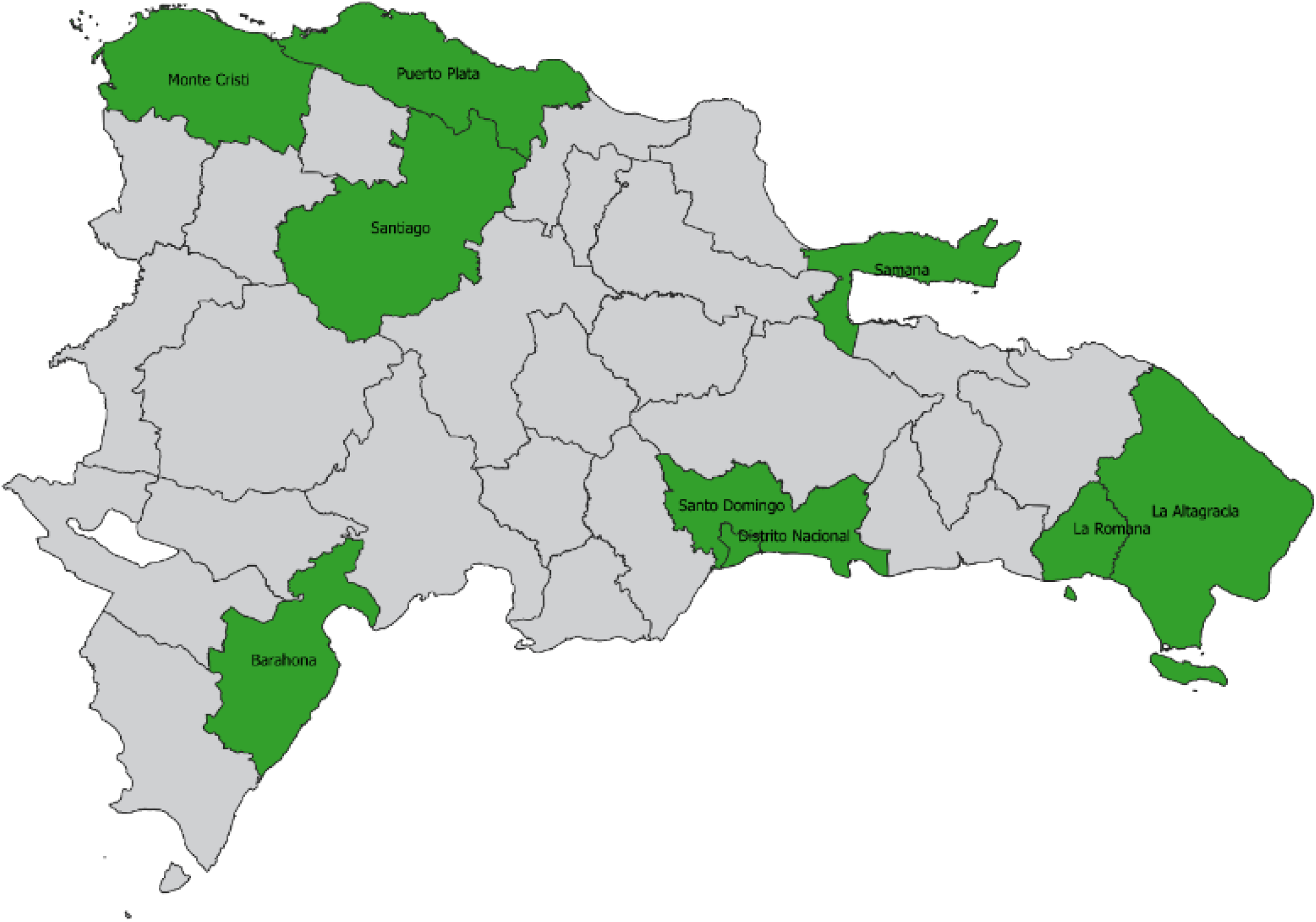
Provinces of the Dominican Republic. Provinces at the focus of this study are highlighted in green.

### 2.2. Data collection

#### Dengue cases

The number of weekly reported cases for the period January 2015 to December was provided by Sistema Nacional de Vigilancia Epidemiológica de la Dirección General de Epidemiológica (Ministerio de Salud Pública). The epidemiological week was defined as Sunday to Saturday. Cases include suspected and laboratory-confirmed cases aggregated at the province level according to surveillance definitions [33]. Dengue Incidence Rate (DIR) was calculated using the number of new cases, divided by the local population each year, multiplied by 100,000 inhabitants. Figure 2a shows dengue incidence for the five years across the nine provinces included in the study.

**Figure 2.**
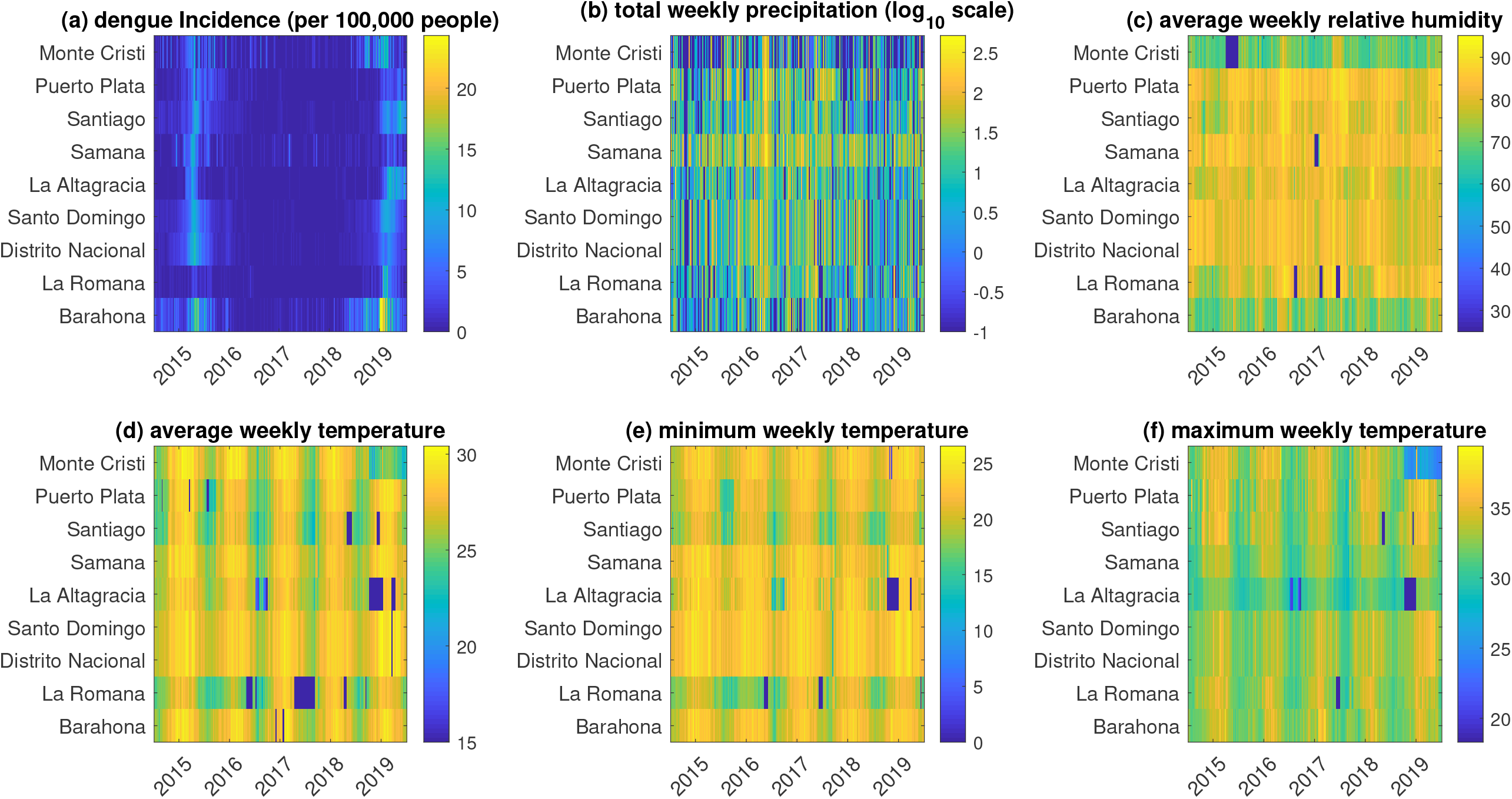
Time series of epidemiological and meteorological metrics across the five years and nine provinces at the focus of this study.

#### Meteorological data

Meteorological data were obtained by supplementing official national data (from ONAMET) with data provided by the U.S. National Aeronautics and Space Administration (NASA). Previous studies have confirmed this approach for collecting data, especially where there are gaps in reliable data [24, 34]. We calculated summary values (minimum, maximum, mean, sum) of meteorological parameters by epidemiologic weeks. Figure 2b,c,d show average weekly temperature, total weekly precipitation, and average weekly relative humidity for the five years across the nine provinces included in the study.

#### Population data

We obtained population data from Oficina Nacional de Estadísticas (ONE) [27]. This data includes the total population and the population density for each province (Table 1). Distrito Nacional has the highest population density, and Santo Domingo province has the highest population.

**Table 1.** Demographic characteristics from provinces of Dominican Republic included in this study, 2019. Data was obtained from ONE [27].

| Province | Population Size | Population density (individuals/km<sup>2</sup>) |
| --- | --- | --- |
| Barahona | 189 149 | 1 160.28 |
| La Altagracia | 345 822 | 114.88 |
| La Romana | 270 166 | 1 456.26 |
| Distrito Nacional | 1 036 494 | 9 924.30 |
| Monte Cristi | 116 605 | 255.99 |
| Puerto Plata | 332 386 | 653.01 |
| Samaná | 111 217 | 130.27 |
| Santiago | 1 038 044 | 369.90 |
| Santo Domingo | 2 855 892 | 1059.03 |

### 2.3. Statistical analysis

#### Summary Statistics

We conducted a preliminary statistical analysis of epidemiological data to highlight changes in dengue cases across provinces and across years. We calculated summary statistics of dengue cases for the entire country each year. We focus much of our analysis of cases on the years 2015 and 2019, when large epidemics took place. Our initial findings support subsequent analysis at the province level to assess the association between explanatory variables and the distribution of the disease over space and time. We calculate dengue incidence in each province in 2015 and 2019 as well as descriptive statistics such as mean, maximum, minimum, and standard deviation of meteorological variables for each province. The results were obtained by calculations in Microsoft Excel, through the Excel Data Analysis Tool. Maps with spatial data were generated by QGIS 3.16.6. These maps improve our understanding of dengue transmission in different regions and how transmission evolves spatially over time [35].

#### Correlated Lags analysis

We assume a unidirectional relationship between dengue cases and meteorological variables. We calculated cross-correlation functions for different weekly summary data for these variables with lags up to 10 weeks. We chose this cutoff for lags because it is a biologically reasonable time for weather events to impact mosquito development and disease transmission. We tested for significance of correlated lags with a two-tailed t-test to test the null hypothesis that the correlation was equivalent to 0. We report the lags with the highest correlation along with p-values at the 0.10, 0.05, and 0.01 confidence levels.

We also conducted a correlated lag analysis among provinces. We calculated correlations between lags in cases in each province. We determined the significance of these correlations with a two-tailed t-test to test the null hypothesis that the correlation was equivalent to 0. We report the lags with the highest correlation along with p-values at the 0.10, 0.05, and 0.01 confidence levels. All correlation analyses were conducted in Matlab 2019a.

## 3. Results

### 3.1. Dengue cases: spatiotemporal analysis

We first calculated descriptive statistics for dengue cases in the country each year (total cases, mean cases per week, standard deviation of cases per week, minimum number of cases per week, maximum number of cases per week, and the dengue incidence rate per year). Table 2 presents these results. In 2015 and 2019 there were major outbreaks with dengue incidence rates of 168.69 and 194.27 cases per 100,000 residents, respectively.

**Table 2.** Descriptive statistics for dengue cases each year, 2015-2020.

|  | 2015 | 2016 | 2017 | 2018 | 2019 | 2020 |
| --- | --- | --- | --- | --- | --- | --- |
| **Total cases** | 16836 | 6559 | 1335 | 1538 | 20123 | 3070 |
| **Mean (per week)** | 323.77 | 126.13 | 25.67 | 29.58 | 386.98 | 59.04 |
| **St. Dev (per week)** | 40.64 | 15.62 | 1.44 | 2.47 | 37.49 | 15.50 |
| **Min (per week)** | 66 | 15 | 5 | 5 | 57 | 0 |
| **Max (per week)** | 1140 | 530 | 57 | 87 | 888 | 417 |
| **DIR (per 100,000 people)** | 168.69 | 65.10 | 13.13 | 14.98 | 194.27 | 29.38 |

The two largest outbreaks each started during late spring (April-May) and continued for approximately one year (Figure 3). The 2015 outbreak peaked in October, while the 2019 outbreak peaked in August. Central provinces experienced the highest incidence during the 2015 outbreaks, while in 2019, provinces in the north and south experienced the highest incidence (Figure 4). The increase in incidence in the northern and southern provinces could be related to socioeconomic factors or differences in climate variables between the two years. Barahona in the southwest, along with Hermanas Mirabel, Sánchez Ramirez, and San José de Ocoa in the center and Hato Mayor in the east all experienced similarly high incidence in both 2015 and 2019. Puerto Plata in the north along with Distrito Nacional in the southeast and many other coastal provinces experienced similarly lower incidence in both outbreaks. Puerto Plata and Distrito Nacional are both popular destinations for international travel and thus are likely to employ more aggressive mosquito control and dengue prevention practices. Table 3 shows incidence calculations for the nine provinces of focus for this study along with the national incidence for both 2015 and 2019.

**Table 3.** – Dengue incidence rate per 100.000 population by province for the 2015 and 2019 outbreaks.

| Year | Barahona | Distrito Nacional | La Altagracia | La Romana | Monte Cristi | Puerto Plata | Samaná | Santiago | Santo Domingo | Dominican Republic |
| --- | --- | --- | --- | --- | --- | --- | --- | --- | --- | --- |
| **2015** | 273.9 | 197.6 | 87.6 | 30.7 | 132.1 | 114.3 | 131.3 | 148.5 | 162.0 | 168.7 |
| **2019** | 456.2 | 163.9 | 206.6 | 153.0 | 250.8 | 104.7 | 72.2 | 216.1 | 210.7 | 194.3 |

**Figure 3.**
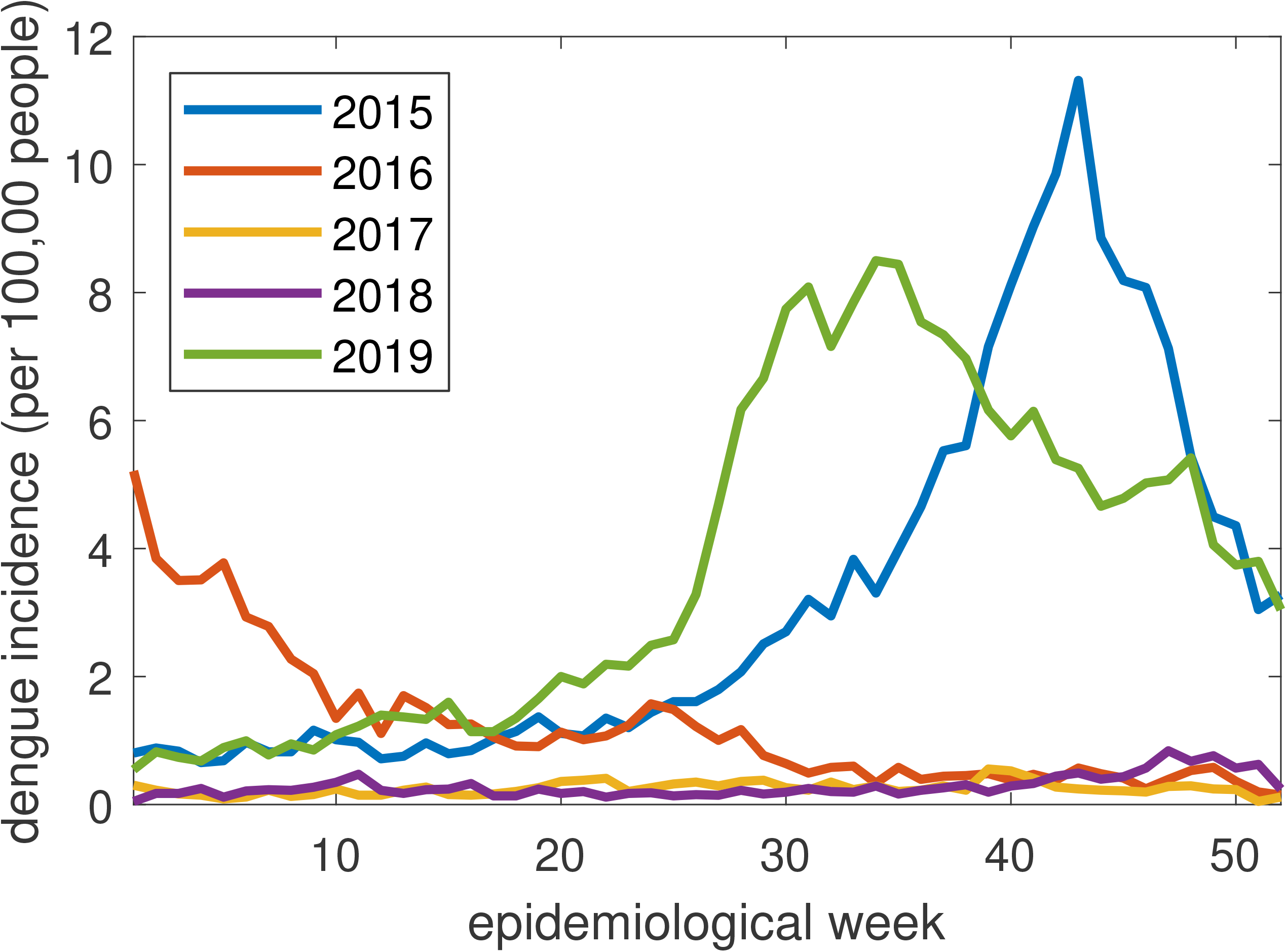
Dengue incidence by week from 2015-2019 in Dominican Republic.

**Figure 4.**
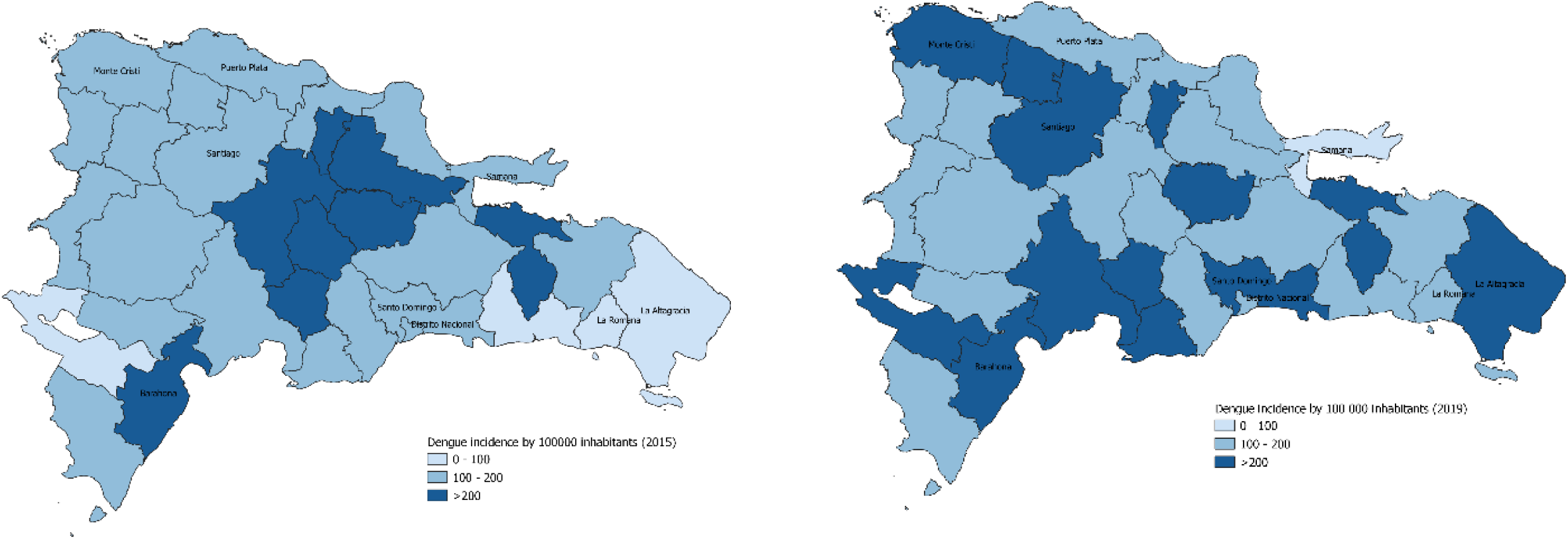
Spatial description of dengue incidence in 2015 (left) and 2019 (right).

### 3.2. Climate variables and dengue cases

The majority of the Dominican Republic has a tropical climate with hot temperatures all year and the warmest months being May to October. There is a rainy season between late April and October, while the northern coast, exposed to the trade winds, is rainy throughout the year. On the southern coast, there is a considerable amount of precipitation because it is not protected by mountains. As a Caribbean country, the rains occur mainly as short showers and thunderstorms which are sometimes intense and often concentrated in short periods of time. Table 4 summarizes the climate statistics for the nine provinces studied in the two outbreaks. In both years, the values for all climate variables did not vary so much.

**Table 4.** Summary statistics of climate variables. Statistics are calculated per week. Averages across the year are shown above ranges (Min-Max) of each variable in parentheses below. All temperatures are given in (°C), and precipitation is given in mm.

|  | 2015 |  |  |  |  | 2019 |  |  |  |  |
| --- | --- | --- | --- | --- | --- | --- | --- | --- | --- | --- |
|  | Min Temp. | Mean Temp. | Max Temp. | Total Precip. | Mean Relative humidity | Min Temp. | Mean Temp. | Max Temp. | Total Precip. | Mean Relative humidity |
| **Santo Domingo** | 23.0<br>(20.8-26) | 27.9<br>(26.0-29.9) | 32.9<br>(31-37.2) | 22.86<br>(0-126.6) | 82.5<br>(72.0-91.5) | 22.9<br>(20.1-26) | 28.3<br>(26.1-30.4) | 33.8<br>(31-37) | 13.7<br>(0-83.8) | 78.6<br>(72.5-84.1) |
| **Barahona** | 21.6<br>(19-25.5) | 27.6<br>(24.5-30) | 33.0<br>(30.4-38) | 9.7<br>(0-79.5) | 69.9<br>(63.3-81.2) | 21.7<br>(18-25) | 27.4<br>(24.9-29.9) | 32.8<br>(30.6-35.8) | 15.0<br>(0-99.6) | 72.5<br>(64.2-83.1) |
| **Distrito Nacional** | 23.0<br>(20.8-26) | 27.9<br>(26.0-29.9) | 32.9<br>(31-37.2) | 22.86<br>(0-126.6) | 82.5<br>(72.0-91.5) | 22.9<br>(20.1-26) | 28.3<br>(26.1-30.4) | 33.8<br>(31-37) | 13.7<br>(0-83.8) | 78.6<br>(72.5-84.1) |
| **La Altagracia** | 22.0<br>(17.1-26.2) | 27.3<br>(25.2-29.4) | 31.4<br>(29.6-33.4) | 18.2<br>(0-272.6) | 78.4<br>(64.8-84.2) | 21.3<br>(17-24) | 26.8<br>(24.9-28.9) | 31.2<br>(28.8-33) | 12.0<br>(0-61.4) | 77.2<br>(71.0-83.2) |
|  | 19.0 | 26.3 | 33.3 | 13.0 | 76.2 | 19.4 | 26.3 | 33.0 | 21.4 | 82.7 |

| La Romana | (15.5-22) | (24.0-28.6) | (31-36) | (0-130.5) | (62.3-84.2) | (14.4-23.2) | (22.9-28.5) | (30.5-35.8) | (0-150.1) | (76.3-90.2) |
| --- | --- | --- | --- | --- | --- | --- | --- | --- | --- | --- |
| Monte Cristi | 21.7<br>(18.5-24.9) | 27.8<br>(23.8-30.3) | 33.8<br>(30.8-37.4) | 10.3<br>(0-165.5) | 68.5<br>(63.2-81.0) | 21.4<br>(2.2-25) | 24.5<br>(20.3-26.9) | 27.3<br>(23-35.9) | 7.6<br>(0-107.5) | 69.3<br>(60.6-77.6) |
| Puerto Plata | 20.8<br>(15.8-23.7) | 27.0<br>(23.6-29.7) | 33.5<br>(30-37.4) | 24.0<br>(0-225.9) | 81.4<br>(68.3-89.0) | 20.9<br>(15.6-23.6) | 27.2<br>(24.2-29.9) | 33.8<br>(30.2-37.7) | 17.6<br>(0-149) | 81.8<br>(77.4-90.5) |
| Samaná | 22.7<br>(29.5-35) | 27.7<br>(25.3-30.2) | 32.4<br>(29.5-35) | 39.4<br>(0-201.9) | 83.6<br>(71.2-91.8) | 22.1<br>(14-25) | 27.6<br>(24.7-29.8) | 32.8<br>(29.5-35) | 30.7<br>(0-148) | 83.2<br>(77.0-89.4) |
| Santiago | 19.6<br>(15-22.8) | 26.6<br>(23.5-29.5) | 33.3<br>(29.7-37.7) | 13.5<br>(0-50.2) | 78.3<br>(65.1-88.0) | 19.2<br>(15-22) | 26.7<br>(23.4-29.3) | 33.8<br>(30.2-37.4) | 16.9<br>(0-140.2) | 79.8<br>(69.7-88.8) |

Although average values and ranges of climate variables are useful for pointing out variations across years, it is important to consider the temporal variation in climate variables and how they might relate to dengue outbreaks. As an example, we show in Figure 5 temporal variation in climate variables and dengue cases in 2015 and 2019 in Distrito Nacional. Trends across provinces were similar and are excluded here for brevity. Temperature and relative humidity are relatively stable throughout the year, although temperatures trend upward from the beginning of each year until epidemiological weeks 30-35. The cumulative precipitation per week is rather variable and could potentially have more influence on dengue cases. We investigate this relationship, along with relationships between weekly variation in other climate variables and dengue transmission, in the next section.

**Figure 5.**
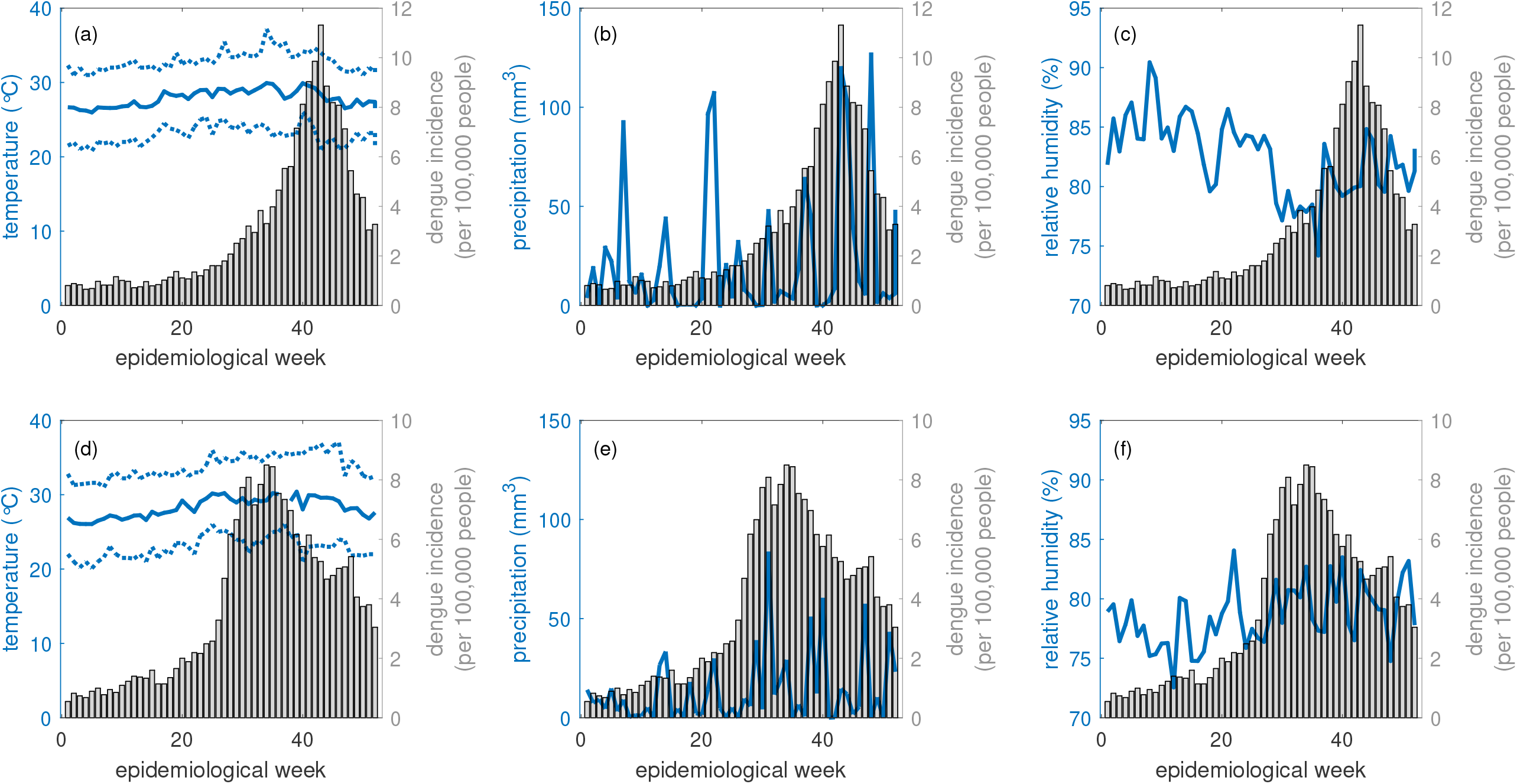
Climate variables and dengue cases in 2015 (a-c) and 2019 (d-f). Variables included are (a,d) temperature (mean temperature is given by the solid curve); (b,e) precipitation; and (c,f) relative humidity.

### 3.3 Cross-correlation analysis

#### Correlations in lags between dengue cases and climate variables

Table 5 contains all correlations between lags in climate variables and dengue cases in the 9 provinces. Lags between dengue cases and temperature variables were strongly correlated among all climate variables we considered. We found the strongest correlations in lags between dengue cases and average weekly temperature, maximum weekly temperature, and minimum weekly temperature. Average temperature between 2 and 10 weeks was strongly positively correlated with dengue in 8 provinces. Maximum temperatures with lags of 4-10 weeks were positively correlated with dengue cases in 8 provinces. Minimum temperatures with lags of 0-10 weeks were positively correlated with dengue cases in all 9 provinces. Notably, in Monte Cristi, average temperature two weeks prior and maximum temperatures 7 weeks prior were negatively correlated with dengue cases, and minimum temperature 0 weeks prior was positively correlated with dengue cases.

**Table 5.** Correlations in lags between dengue cases and climate variables. Lags are given as the number of weeks prior to dengue cases. Lags are listed with correlations in parentheses. Stars indicate confidence levels for testing significance: *** p<.01, ** p< .05, *p <.10.

| PROVINCE | Mean Daily Temperature | Mean Daily Temperature Range | Max. Weekly Temperature | Min. Weekly Temperature | Mean Daily Precipitation | Total Weekly Precipitation | Mean Daily Relative Humidity | Max. Weekly Relative Humidity | Min. Weekly Relative Humidity |
| --- | --- | --- | --- | --- | --- | --- | --- | --- | --- |
| Barahona | -9<br>(0.1908)*** | 0<br>(-0.0461) | -7<br>(0.1507)** | -10<br>(0.2269)*** | -5<br>(-0.0557) | -5<br>(-0.0580) | -2<br>(0.0534) | -6<br>(0.0549) | -2<br>(0.0688) |
| La Altagracia | -10<br>(0.3300)*** | -2<br>(0.1877)*** | -5<br>(0.3278)*** | -9<br>(0.2645)*** | -1<br>(-0.0520) | -1<br>(-0.0400) | -3<br>(0.0535) | 0<br>(0.0638) | -1<br>(0.0770) |
| La Romana | -4<br>(0.2765)*** | -4<br>(-0.0733) | -4<br>(0.2487)*** | -6<br>(0.2428)*** | -2<br>(0.0371) | -2<br>(0.0347) | 0<br>(0.1348)** | 0<br>(0.1143)* | 0<br>(0.0947) |
| Monte Cristi | -2<br>(-0.1920)*** | -10<br>(-0.2988)*** | -7<br>(-0.2632)*** | 0<br>(0.0895) | -3<br>(-0.0235) | -3<br>(-0.0234) | -1<br>(-0.0495) | -1<br>(-0.0492) | -9<br>(0.0513) |
| Puerto Plata | -9<br>(0.3081)*** | -10<br>(0.2270)*** | -10<br>(0.3555)*** | -9<br>(0.1992)*** | 0<br>(-0.0505) | 0<br>(-0.0525) | 0<br>(-0.1015) | 0<br>(-0.0775) | -1<br>(-0.0432) |
| Samaná | -7<br>(0.3662)*** | 0<br>(0.1055)* | -7<br>(0.3335)*** | -5<br>(0.3092)*** | -4<br>(-0.0500) | -4<br>(-0.0541) | -5<br>(0.0788) | 0<br>(0.0706) | -4<br>(0.0704) |
| Santiago | -10<br>(0.4408)*** | -10<br>(0.3180)*** | -10<br>(0.4667)*** | -10<br>(0.2833)*** | -8<br>(-0.0576) | -8<br>(-0.0509) | 0<br>(-0.0799) | 0<br>(-0.0865) | 0<br>(-0.0414) |
| Santo Domingo | -5<br>(0.4384)*** | 0<br>(0.1424)** | -4<br>(0.3941)*** | -5<br>(0.3028)*** | 0<br>(-0.0609) | 0<br>(-0.0664) | 0<br>(-0.2641)*** | 0<br>(-0.2461)*** | 0<br>(-0.1148)* |
| Distrito Nacional | -7<br>(0.3982)*** | 0<br>(0.0944) | -6<br>(0.3429)*** | -9<br>(0.2895)*** | -1<br>(-0.0496) | 0<br>(-0.0577) | -1<br>(-0.2470)*** | -1<br>(-0.2438)*** | -1<br>(-0.0981) |

Lags between mean daily average temperature and dengue cases were also significantly correlated in some provinces. In La Altagracia, Puerto Plata, and Santiago, these correlations were positive at 2, 10, and 10 weeks, respectively; however, in Monte Cristi, the correlation was negative at 10 weeks. In Santo Domingo and Distrito Nacional, we found significant correlations between average and maximum humidity and dengue cases, but no humidity variables were significantly correlated with dengue cases in any other provinces.

No correlations of lags with total precipitation and average precipitation were significant at the α=.05 confidence level. This result is surprising because we would expect dengue transmission in tropical climates to be positively correlated with precipitation given the important role of water in the mosquito’s life cycle [36]

#### Correlations in lags between cases in different provinces

All the largest correlations in lags between cases in different provinces were highly significant (p<.01). In most cases, correlations between provinces with a lag of τ=0 were the strongest of all of the lags, suggesting that cases typically occurred simultaneously across provinces (Table 6). Cases in Distrito Nacional were generally in sync with cases in other regions, with cases in Distrito Nacional with a lag of τ=0 having strong positive correlations with cases in all other provinces except Monte Cristi (τ=-3), and cases in Monte Cristi with a lag of τ=0 were strongly positively correlated with cases in all other provinces except La Romana (τ=-1). In Barahona, cases with a lag of τ=0 were strongly positively correlated with cases in all other provinces except Samaná (τ=-3). In each of these, the non-zero lag is in a province that is typically far away from the other province. For example, Barahona and Samaná are on opposite sides of the country, as are Monte Cristi and Distrito Nacional.

**Table 6.** Correlations in lags (weeks) between dengue cases in each province. In this table, the provinces along the columns are the predictor variables. Lags are listed with correlations in parentheses. Stars indicate confidence levels for testing significance: *** p<.01, ** p< .05, *p <.10.

| PROVINCE | Barahona | La Altagracia | La Romana | Monte Cristi | Puerto Plata | Samaná | Santiago | Santo Domingo | Distrito Nacional |
| --- | --- | --- | --- | --- | --- | --- | --- | --- | --- |
| Barahona |  | 0<br>(0.4751)*** | 0<br>(0.6413)*** | 0<br>(0.6741)*** | 0<br>(0.5285)*** | -3<br>(0.4404)*** | 0<br>(0.6002)*** | 0<br>(0.8161)*** | 0<br>(0.7509)*** |
| La Altagracia | -8<br>(0.6177)*** |  | -7<br>(0.7454)*** | -6<br>(0.5034)*** | 0<br>(0.6211)*** | 0<br>(0.5462)*** | 0<br>(0.8134)*** | -2<br>(0.7693)*** | 0<br>(0.6680)*** |
| La Romana | -3<br>(0.7418)*** | 0<br>(0.5840)*** |  | -1<br>(0.5741)*** | 0<br>(0.3917)*** | 0<br>(0.2985)*** | 0<br>(0.5875)*** | 0<br>(0.7421)*** | 0<br>(0.5661)*** |
| Monte Cristi | 0<br>(0.6741)*** | 0<br>(0.3616)*** | -1<br>(0.5579)*** |  | 0<br>(0.4680)*** | 0<br>(0.3918)*** | 0<br>(0.4656)*** | 0<br>(0.6483)*** | 0<br>(0.5831)*** |
| Puerto Plata | -6<br>(0.5444)*** | -2<br>(0.6405)*** | -1<br>(0.4495)*** | -6<br>(0.4885)*** |  | 0<br>(0.6544)*** | 0<br>(0.7985)*** | -4<br>(0.7185)*** | 0<br>(0.7557)*** |
| Samaná | -2<br>(0.4070)*** | 0<br>(0.5462)*** | -1<br>(0.3079)*** | 0<br>(0.3918)*** | 0<br>(0.6544)*** |  | 0<br>(0.5770)*** | 0<br>(0.6385)*** | 0<br>(0.7162)*** |
| Santiago | -4<br>(0.6413)*** | -3<br>(0.8420)*** | -2<br>(0.6100)*** | -7<br>(0.5238)*** | 0<br>(0.7985)*** | -2<br>(0.5796)*** |  | -4<br>(0.8297)*** | -4<br>(0.7749)*** |
| Santo Domingo | -1<br>(0.8242)*** | 0<br>(0.7413)*** | 0<br>(0.7421)*** | -4<br>(0.6656)*** | 0<br>(0.7075)*** | 0<br>(0.6385)*** | 0<br>(0.7929)*** |  | 0<br>(0.9364)*** |
| Distrito Nacional | 0<br>(0.7509)*** | 0<br>(0.6680)*** | 0<br>(0.5661)*** | -3<br>(0.6228)*** | 0<br>(0.7557)*** | 0<br>(0.7162)*** | 0<br>(0.7566)*** | 0<br>(0.9364)*** |  |

In Santo Domingo, Samaná, and La Romana, the results were similar: in each of these three provinces correlations in cases with a lag of τ=0 were strongly positively correlated with cases in all other provinces except two. For Santo Domingo, the exceptions were Barahona (τ=-1) and Monte Cristi (τ=-4). For Samaná, the exceptions were Barahona (τ=-2) and La Romana (τ=-1). For La Romana, the exceptions were Barahona (τ=-3) and Monte Cristi (τ=-1). Again, the provinces in these exceptions are geographically distant from the provinces whose cases follow 1-4 weeks later.

La Altagracia, Puerto Plata, and Santiago had more non-zero lags that were significant. Cases in La Altagracia were positively correlated with cases in Barahona, La Romana, Monte Cristi, and Santo Domingo that occurred 8, 7, 6, and 2 weeks prior, respectively. Cases in Puerto Plata were positively correlated with cases in Barahona (τ=-6), La Altagracia (τ=-2), La Romana (τ=-1), Monte Cristi (τ=-6), and Santo Domingo (τ=-2). In Samaná, cases were positively correlated with cases in 7 of the 8 other provinces: Barahona (τ= -4), La Altagracia (τ= -3), Monte Cristi (τ=-7), Samaná(τ =-2), Santo Domingo (τ=-4), and Distrito Nacional (τ= -4).

## 4. Discussion

Herein we characterized dengue incidence at the province level in Dominican Republic between 2015-2019, a period in which the country and the Caribbean region experienced two large epidemics. We focused our study on nine provinces that included all major geographic regions of the country that represented different climate patterns. In our study, we observed different potential drivers of dengue activity in different regions of the country. We anticipate that this study will be a foundation upon which models aimed at predicting dengue activity may be built.

We noted that both major outbreaks (2015 and 2019) occurred after the 30_th_ epidemiological week, which generally corresponds approximately to late July. This, together with the fluctuations noted even in the years in which no epidemic occurred, indicate a seasonal pattern of dengue transmission. When comparing the epidemiology of dengue in the country with the region of the Americas, a similar behavior was observed for 2015 and 2019, the latter being the year with the highest number of cases recorded in the history of dengue in the Americas [28, 37]. The reduction in the number of cases between 2016-2018 could be explained in part by the vector control actions implemented by the Ministry of Health, the adaptation of the pathogen, reduction of susceptible population, or partial immunity to dengue conferred by the wave of Zika virus that moved through the region between 2015-2016 [38, 39].

We find that the southwestern province of Barahona had the largest dengue incidence in both 2015 (273.9 per 100,000 people) and 2019 (456.2). Furthermore, dengue activity in Barahona preceded dengue activity in most other provinces by up to eight weeks. It is possible that new cases are introduced to the Dominican Republic in this region through immigration from Haiti or via tourism. Cases in Monte Cristi, too, preceded cases elsewhere in the country by 1-7 weeks. It is possible that for both provinces, individuals who have dengue must travel to other provinces for medical care as both provinces only have one public hospital[40]. This could lead to movement of cases into other provinces and throughout the country.

Our analyses of lags in cases between provinces could help determine how cases spread spatially in the country by identifying “source” provinces where dengue cases begin (such as Barahona and Monte Cristi) and “sink” provinces where dengue cases later appear. For example, cases in La Altagracia, Puerto Plata, and Santiago often trailed cases in other parts of the country. Both La Altagracia and Puerto Plata are home to several popular tourist attractions and may benefit from increased surveillance and vector control [41]. It is possible that as cases are reported elsewhere in the country, control efforts delay significant amounts of transmission in these provinces. Despite these lags found, most of the lags between provinces were zero, indicating that outbreaks occurred throughout most of the region at about the same time. This could be explained by human movement inside the country. A similar study in the country with Zika virus also suggested that the human mobility and the infrastructure level of each region could influence the transmission of diseases that had *Aedes* aegypti as a vector [42].

We analyzed climate variables that could contribute to the dengue transmission cycle by their impacts on the *Ae. aegypti* life cycle. In Santo Domingo, the temperature and relative humidity do not vary much throughout the year. However, there are some weeks where the rainfall was more intense and could contribute to additional hatching of mosquito eggs. However, when all five years of dengue case and climate data are considered, lags between temperature variables and dengue cases were most highly correlated, indicating temperature as a good predictor of dengue transmission throughout the region. This result is supported by work showing that temperature influences dengue transmission through its impacts on both the vector life cycle and the virus [15, 19, 43, 44]. Surprisingly, lags between precipitation and dengue cases were not found to be significantly correlated with cases in any of the provinces. In studies of other tropical regions, precipitation and humidity are often found to be positively correlated with arbovirus activity [29, 45, 46]. It is possible that this is because Dominican Republic’s unique topography interferes with weather patterns and results in having rainy seasons at different times of the year in different regions [29]. While cases could be impacted locally by changes in precipitation, this may not correspond to times at which dengue transmission is occurring elsewhere, which may lead to impacts on correlation. These results are in line with the ones achieved in [47], showing that temperature and humidity have impact in the transmission chain.

## 5. Conclusions

The short period of data included in this study is insufficient for making strong characterizations of relationships. However, in this work we developed a better understanding of which variables have been most strongly associated with dengue cases in this time frame, which includes two large outbreaks. These findings will help inform future work for building predictive models that incorporate climate and spatiotemporal data to characterize province risk and refine public health responses. This study contributes an important analysis of recent dengue transmission on which more complex spatiotemporal analyses can be conducted. The general characterizations of climate and dengue activity along with the correlated lags analysis across the nine provinces included here provide a foundation upon which future studies may build to investigate more intricate relationships between dengue and climate, human movement, and human activity.

## Data Availability

The datasets analyzed during the current study are available from the corresponding author on request. Upon publication, data will be publicly available in a GitHub repository.

## 7. Acknowledgements

The authors would like to thank Albert Rodriguez, Heyliana Marte and Pedro Vegas for their contributions to this project.

## 9. Ethics Approval

Not applicable.

## 10. Consent for Publication

Not applicable.

## 13. Author’s Contributions

MAR, HSR, DH, and MC-H conceived and designed the study. MAR and HSR conducted statistical analyses. MAR and HSR drafted the manuscript. All authors contributed to interpretation of the data and revisions of the manuscript. All authors read and approved the final manuscript.

